# Protocol of the Can Nebulised Heparin Reduce Time to Extubation in SARS-CoV-2 (CHARTER) Study

**DOI:** 10.1101/2020.04.28.20082552

**Authors:** Barry Dixon, Roger J Smith, Antonio Artigas, John Laffey, Bairbre McNicholas, Eric Schmidt, Quentin Nunes, Mark Skidmore, Marcelo Andrade de Lima, John L Moran, Frank VanHaren, Gordon Doig, Angajendra Ghosh, Simone Said, Sachin Gupta, John D Santamaria

## Abstract

**Introduction:** COVID-19 is associated with the development of ARDS displaying the typical features of diffuse alveolar damage with extensive pulmonary coagulation activation resulting in fibrin deposition in the microvasculature and formation of hyaline membranes in the air sacs. The anti-coagulant actions of nebulised heparin limit fibrin deposition and progression of lung injury. Serendipitously, unfractionated heparin also inactivates the SARS-CoV-2 virus and prevents its entry into mammalian cells. Nebulisation of heparin may therefore limit both fibrin-mediated lung injury and inhibit pulmonary infection by SARS-CoV-2. For these reasons we have initiated a multi-centre international trial of nebulised heparin in patients with COVID-19.

**Methods and intervention:** Mechanically ventilated patients with confirmed or strongly suspected SARS-CoV-2 infection, hypoxaemia and an acute pulmonary opacity in at least one lung quadrant on chest X-ray, will be randomised to nebulised heparin 25,000 Units every 6 hours or standard care for up to 10 days while mechanically ventilated. The primary outcome is the time to separation from invasive ventilation to day 28, where non-survivors to day 28 are treated as though not separated from invasive ventilation.

**Ethics and dissemination:** The study protocol has been submitted to the human research and ethics committee of St Vincent’s Hospital, Melbourne, Australia. Submission is pending in other jurisdictions. Results of this study will be published in scientific journals and presented at scientific meetings.

**Trial Registration:** ACTRN: 12620000517976

## Introduction

A recent (not yet published; under journal review) pre-pandemic double-blind multi-centre randomised study of 256 mechanically ventilated patients with or at risk of developing ARDS led by our group found, clinically important pre-specified secondary outcomes were significantly improved with nebulised heparin. There was no evidence of harm.

COVID-19 is associated with the development of ARDS displaying the typical features of diffuse alveolar damage with extensive pulmonary coagulation activation resulting in fibrin deposition in the microvasculature and formation of hyaline membranes in the air sacs. The anticoagulant actions of nebulised heparin limit fibrin deposition. Serendipitously, unfractionated heparin also inactivates the SARS-CoV-2 virus and prevents its entry into mammalian cells. Nebulisation of heparin may therefore limit fibrin-mediated lung injury and inhibit pulmonary infection by SARS-CoV-2. For these reasons we believe a trial of nebulised heparin in patients with COVID-19 is warranted.

## Morbidity and mortality of ARDS and COVID-19

ARDS affects 23% of mechanically ventilated critically ill patients and has a mortality up to 46%.^1,2^ The death rate in COVID-19 patients with ARDS may be higher, up to 66%.^3^ Survivors of ARDS have marked limitations in physical function with increased need for rehabilitation and long-term care.^4,5^

## Histological features of ARDS

The clinical syndrome of ARDS due to acute inflammatory changes in the lungs was first described in a 12-patient case series by Ashbaugh et al in 1967.^6^ The histological changes found by Ashbaugh have been termed diffuse alveolar damage.^7^ The hallmark histological feature of ARDS is a fibrin mesh in the air sacs, known as a hyaline membrane, on which leucocytes attach and manifest the inflammatory responses that result in diffuse alveolar damage.^7-9^ Fibrin accumulation in pulmonary capillaries and venules, which leads to microvascular thrombosis is another early feature,^10-12^ and the extent of this fibrin accumulation correlates with the severity of acute lung injury.^11,13^ Many conditions can trigger an inflammatory response that results in diffuse alveolar damage including pneumonia, sepsis, aspiration, transfusion, cardiac surgery and trauma.^14^

## Pulmonary microvascular thrombosis

In response to inflammatory cytokines the pulmonary capillary beds, venules and arterioles express tissue factor on endothelial cells and this triggers the conversion of plasma coagulation factors to fibrin.^15^ Extensive microvascular thrombosis has been demonstrated in histological studies of ARDS.^10,11^ Angiographic studies showed the extent of microvascular obstruction correlated with the severity of respiratory failure and with mortality.^11,13^ Microvascular thrombosis increases lung dead space and the increase in the dead space was shown to be an independent marker of mortality in ARDS.^16^ Microvascular thrombosis also causes increased pulmonary vascular resistance, which may result in right heart failure.^17^

## Hyaline membrane formation

Hyaline membrane formation is a consistent and early manifestation of the inflammatory response in ARDS.^6,10,18-20^ Hyaline membrane formation results from entry into the alveolar space of inflammatory exudate that is rich in plasma borne coagulation factors. The subsequent expression of tissue factor by alveolar epithelial cells and macrophages triggers the conversion of these coagulation factors to fibrin and the formation of the hyaline membrane. The concurrent expression of plasminogen activator inhibitor-1 by alveolar macrophages prevents the removal of this membrane through endogenous fibrinolysis.^8^

Hyaline membrane formation may contribute to lung injury through a number of mechanisms. Fibrin is a pro-inflammatory molecule attracting leucocytes which attach to fibrin promoting further inflammatory responses that results in diffuse alveolar damage.^7-9^ The hyaline membrane forms a physical barrier thereby limiting the diffusion of gases. Alveolar compliance and the action of surfactant is also limited contributing to atelectasis. Finally, the laying down of a fibrin matrix may promote subsequent lung fibrosis.^8,21^

## SARS-CoV-2 is associated with increased inflammatory cytokines and excessive coagulation activation and fibrin deposition in the lungs and other organs

Patients with SARS-CoV-2 who manifest severe disease including ARDS, multi-organ failure and death, have higher levels of inflammatory cytokines (“cytokine storm”), higher plasma markers of coagulation, such as D-Dimers, increased prothrombin time and a lower platelet count.^3,22-25^ Furthermore, multivariate regression found a D-dimer level (>1 mg/L) was an independent risk factor for mortality.^3,26^ Finally, more than 70% of non-survivors developed DIC 2728 Post-mortem studies and lung biopsies of SARS-CoV-2 patients with ARDS demonstrated pulmonary fibrin deposition with hyaline membranes in the alveolar spaces and extensive pulmonary microvascular thrombi.^29-36^ Microvascular thrombi were also found in other organs including the heart, liver and kidneys.^32,34^

## Treatment of pulmonary microvascular thrombosis and hyaline membranes

Nebulised heparin targets pulmonary fibrin deposition. Heparin’s anti-coagulant properties have been used in clinical practice to limit fibrin deposition since 1935. Heparin inhibits coagulation activation through a range of mechanisms, including catalysing the action of antithrombin, promoting tissue factor pathway inhibitor expression, reducing tissue factor expression, increasing endothelial expression of heparan sulphate and through release of t-PA by the endothelium.^37-40^ Heparin also has other actions of potential benefit including inhibition of inflammatory cytokines, prevention of bronchospasm, increased nitric oxide release and limiting adhesion of microbes to respiratory epithelium.^41-44^

Early-phase trials in patients with acute lung injury and related conditions found heparin reduced pulmonary dead space, coagulation activation, microvascular thrombosis and deterioration in the Murray Lung Injury Score and increased time free of ventilatory support.^45-50^

Our recent multi-centre randomised double-blind placebo-controlled trial of nebulised heparin (currently under consideration for publication) in patients with or at risk of developing Acute Respiratory Distress Syndrome, to determine if nebulised heparin improves long term physical function (ANZCTR, 12612000418875), found that amongst 256 mechanically ventilated patients, 47% of whom had ARDS, clinically important prespecified secondary outcomes were significantly improved. There was no evidence of harm. Our results suggest that nebulised heparin limits pulmonary fibrin deposition, attenuates progression of acute lung injury and hastens recovery.

## Heparin administration may be associated with improved clinical outcomes in SARS-CoV-2

Anecdotal data suggest patients with SARS-CoV-2 treated with systemic unfractionated heparin or LMWH had better clinical outcomes.^51^ A non-randomised study found patients with a sepsis-induced coagulation score greater than 4 were more likely to survive if administered heparin or LMWH.^25^

## SARS-CoV-2 inactivation by heparin

Heparin is a member of a family of proteoglycan molecules that include heparan sulphate, chondroitin sulphate, keratan sulphate and hyaluronic acid. These molecules are expressed throughout the body, with diverse biological roles and are usually associated with respiratory and endothelial cell surfaces, basement membrane and extracellular matrices.^52^ In humans heparin is produced solely by mast cells and is stored in granules. Heparin makes up 30% of the dry weight of mast cell granules.^53^

There is evidence that heparin plays a role in host defence. Firstly, mast cells are mostly located along blood vessels and are particularly associated with capillaries and post-capillary venules.^54^ Secondly, organs exposed to the external environment, such as the lungs and gut contain a large proportion of the body’s mast cells.^55^ Thirdly, heparin is conserved across a variety of different species, some of which do not have a blood coagulation system like ours (e.g. molluscs), suggesting heparin has significant biological roles unrelated to coagulation.^56^ Studies in humans have also demonstrated that heparin has a complex range of biological actions independent of coagulation, some of which may also be beneficial in limiting lung damage associated with critical illness.

A large number of bacterial and viral pathogens depend upon interactions with proteoglycan molecules such as heparan sulphate, which is expressed on a range of human tissue surfaces, for adhesion and invasion of host tissues.^52^ A number of studies found heparin competes with heparan sulphate for bacterial and viral adhesion and may therefore limit pathogen invasion.^57,58^ Heparin limits adhesion of a number of pathogens including Pseudomonas aeruginosa, Burkholderia cenocepacia, Burkholderia pseudomallei, Legionella pneumophila, Staph aureus, Strep pyogenes, Strep pneumonia, Respiratory syncytial virus and Influenza a.^44,59-62^ Human and animal studies suggest these actions may reduce the development of pneumonia and bacteraemia.^57,63^

## Unfractionated heparin binds SARS-CoV-2 and prevents cell invasion by Coronavirus

A recent study demonstrated that the SARS-CoV-2 Spike S1 protein receptor binding domain attaches to unfractionated heparin and undergoes conformational change as a result.^64^ Previous studies demonstrated that unfractionated heparin prevented SARS-associated coronavirus and other coronavirus strains from invading mammalian cells.^65-70^

### Safety and tolerability of nebulised heparin

APTT. Treatment with nebulised heparin at the dose proposed for this study, while concomitantly receiving intravenous or subcutaneous unfractionated heparin, is likely to cause a mild (approximately 7 seconds) increase in the average APTT and a mild-moderate (approximately 18 seconds) increase in the peak APTT during the treatment period. The APTT of patients who are not receiving intravenous or subcutaneous unfractionated heparin is likely to be not affected by treatment with nebulised heparin.^71^

Bleeding and blood transfusion. Nebulised heparin at the dose proposed in this study has not been found to increase the risk of major non-pulmonary bleeding or of blood transfusion.^71^ In clinical trials where more than 150 intensive care patients were treated with nebulised heparin for ARDS and related conditions, there were no cases of pulmonary haemorrhage with patient deterioration.^45,49,71,72^ Bloodstaining of the airway secretions of invasively ventilated intensive care unit patients is common. Approximately 1 out of 20 patients treated with nebulised heparin experience greater-than-usual bloodstaining of secretions and, although this bloodstaining can be unsightly, rarely is it clinically deleterious. The risk of medically important haemoptysis is estimated to be less than 1 out of 100.

Use in asthma and COPD. Use in asthma and COPD is typically well-tolerated but, on rare occasions in patients with severe bronchospasm, airflow may be further impeded immediately following the start of nebulisation.^71^ This may represent heparin accumulating on the luminal surface of terminal bronchioles, causing further narrowing. This typically resolves within minutes of stopping nebulisation and withholding the remainder of the dose. Subsequent doses are typically well-tolerated, but vigilance is required.

Heparin induced thrombocytopenia (HIT). HIT is an immune-mediated adverse reaction caused by the emergence of antibodies that activate platelets in the presence of heparin. Patients in intensive care are commonly treated with subcutaneous heparin for the prevention of deep vein thrombosis. The risk of HIT from treatment with subcutaneous unfractionated is less than 1 out of 100. Nebulised heparin is not thought to increase the risk of HIT in patients already receiving unfractionated heparin and administration by nebulisation is not thought to increase the risk of HIT compared to administration by other routes and no patient to date in our earlier trials developed hit.^45,49,71,72^

Clinician exposure to SARS-CoV-2. Administering nebulised heparin will not significantly increase the risk of SARS-CoV-2 exposure for clinicians who follow the standard recommended personal protection precautions for the care of confirmed or suspected COVID-19 cases. The Aerogen vibrating mesh membrane system has an in-line circuit design so the ventilator circuit does not need to be broken for drug delivery. The medication reservoir is isolated from the breathing circuit and is positioned in the inspiratory limb of the ventilator circuit on the ‘dry’ side of the humidifier, remote from the patient’s endotracheal tube. Once the nebuliser is in position it does not need to be removed until the course of treatment is finished.^73^ An expiratory filter is positioned at the end of the expiratory limb of the ventilator circuit. The main purpose of the filter is to prevent exhaled heparin from depositing on the ventilator’s expiratory sensors and valves, but it is also a highly efficient viral filter that will mitigate against entry of exhaled virus into the room.^74^

## Trial Design and Feasibility

This is an investigator-initiated, multi-centre, randomised, open-label trial of nebulised heparin sodium in addition to standard care compared to standard care alone in 172 invasively ventilated ICU patients with suspected or confirmed COVID-19 infection. Nebulised heparin is administered 6-hourly to day 10 while the patient is receiving invasive mechanical ventilation.

There is likely to be considerable variation in the number of severe COVID-19 infections in different regions. However, the majority of the COVID-19 patients requiring invasive ventilation in intensive care are likely to satisfy the study eligibility criteria, and the investigators, having conducted several clinical trials of nebulised heparin and having established links within the intensive care community, are in an excellent position to identify and promptly resolve potential threats to the successful conduct of the trial.

## Assessment of Efficacy, Safety and Processes-of-Care

### Primary outcome

The primary outcome is the time to separation from invasive ventilation to day 28, where non-survivors to day 28 are treated as though not separated from invasive ventilation. Previous studies have shown that among critically ill patients with ARDS and related conditions, nebulised heparin reduces the duration of mechanical ventilation.^47^

‘Invasive ventilation’ means any positive pressure ventilatory support via an endotracheal or tracheostomy tube. If a patient achieves separation from invasive ventilation more than once, it is the final separation that is used to calculate the outcome.

In this study, ‘day 0’ describes the period from randomisation to midnight on the day of enrolment, ‘day 1’ the first calendar day after the day of enrolment, ‘day 2’ the second calendar day after the day of enrolment, and so forth.

### Secondary outcomes

- Change in oxygenation index, driving pressure and ventilatory ratio at day 2
- Change in white cell count, platelet count, C-reactive protein, D-dimer and INR to day 10
- Number treated with neuromuscular blockers instituted after enrolment to day 10
- Number treated with prone positioning instituted after enrolment to day 10
- Number treated with ECMO instituted after enrolment to day 10
- Number tracheotomised to day 28
- Time to separation from invasive ventilation to day 28, among survivors
- Time to separation from the ICU to day 28, where non-survivors to day 28 are treated as though not separated from invasive care
- Time to separation from the ICU to day 28, among survivors During the pandemic critically ill inpatients might be cared for outside of the walls of the usual physical environment of ICU. For this reason, ‘ICU’ is defined as an area designated for inpatient care of the critically ill where therapies including invasive mechanical ventilation can be provided.
- Survival to day 28; Survival to day 60; and Survival to hospital discharge, censored at day 60
- Number residing at home or in a community setting at day 60
- Number residing at home or in a community setting at day 60, among survivors

### Safety outcomes

- Change in haemoglobin to day 10
- Number transfused red blood cells (packed red cells and whole blood) to day 10
- Volume of red blood cells (packed red cells and whole blood) transfused to day 10
- Number who record major bleeding ‘Major bleeding’ is defined as: bleeding that results in death and/or; bleeding that is symptomatic and occurs in a critical area or organ (intra-cranial, intra-spinal, intra-ocular, retroperitoneal, intra-articular, or intramuscular with compartment syndrome) and /or; bleeding that results in a fall in haemoglobin of 20g/L or more, or results in transfusion of two or more units of whole blood or red cells.^77^
- Number who record pulmonary bleeding ‘Pulmonary bleeding’ is frank bleeding in the lungs, trachea or bronchi with repeated haemoptysis or requiring repeated suctioning <u>and</u> associated with acute deterioration in respiratory status.
- Number who record heparin-induced thrombocytopaenia (HIT) ‘HIT’ is an unexplained fall in platelet count <u>and</u> a positive heparin antibody test.
- Number who record other adverse events and reactions ‘Adverse events and reactions’ are those that, in the site Principal Investigator’s judgement, are not part of the expected clinical course <u>and</u> could be related (at least possibly) to the study <u>and</u> were medically significant or had serious sequelae for the patient.

### Process of care assessments

- Time from intubation to randomisation
- Total cumulative dose of nebulised heparin in ICU to day 10
- Days of treatment with nebulised heparin in ICU to day 10
- Mean APTT in ICU to day 10 among all participants, and among those treated with intravenous or subcutaneous unfractionated heparin, and among those not treated with intravenous or subcutaneous unfractionated heparin
- Highest APTT in ICU to day 10 among all participants, and among those treated with intravenous or subcutaneous unfractionated heparin, and among those not treated with intravenous or subcutaneous unfractionated heparin
- Days of treatment with each of the following therapies while in ICU to day 10: unfractionated heparin, intravenous and subcutaneous; LMWH, intravenous and subcutaneous; lopinavir-ritonavir; remdesivir; hydroxychloroquine; interferonβ; interleukin antagonists; oseltamivir, laninamivir, zaninamivir or peramivir; macrolide; non-macrolide antibacterial; antifungal; corticosteroid; inotrope or vasopressor infusion; and renal replacement.

## Eligibility

### Inclusion criteria

To be eligible, a patient must satisfy <u>all</u> these inclusion criteria:

- Confirmed or strongly suspected COVID-19 Results must be pending or testing planned if COVID-19 is ‘suspected’
- Age 18 years or older
- Endotracheal tube in place
- Intubated yesterday or today
- P_a_O_2_ to F_I_O_2_ ratio less than or equal to 300 while intubated
- Acute opacities not fully explained by effusions, lobar/lung collapse and nodules, affecting at least one lung quadrant on chest X-ray or CT
- Currently in the ICU or scheduled for transfer to the ICU. The ‘ICU’ is an area designated for inpatient care of the critically ill where therapies including invasive mechanical ventilation can be provided.

### Exclusion criteria

To be eligible, a patient must have <u>none</u> of these exclusion criteria:

- Enrolled in another clinical trial that is unapproved for co-enrolment*
- Heparin allergy or HIT
- APTT > 120 seconds and this is not due to anticoagulant therapy
- Platelet count < 20 x 10^9^ per L
- Pulmonary bleeding, which is frank bleeding in the trachea, bronchi or lungs with repeated haemoptysis or requiring repeated suctioning
- Uncontrolled bleeding
- Pregnant or might be pregnant Females aged 18-49 years are excluded unless there is documented menopause or hysterectomy or a pregnancy test was performed and is negative.
- Receiving or about to commence ECMO or HFOV
- Myopathy, spinal cord injury, or nerve injury or disease with a likely prolonged incapacity to breathe independently e.g. Guillain-Barre syndrome
- Usually receives home oxygen
- Dependent on others for personal care due to physical or cognitive decline
- Death is imminent or inevitable within 24 hours
- The clinical team would not be able to set up the study nebuliser and ventilator circuit as required including with active humidification
- Clinician objection.

### Screening Procedures

Research coordinators and investigators at each site will work with clinicians to identify potential candidates for enrolment. A log will be maintained of patients who met the inclusion criteria but were not enrolled, with the reason for exclusion recorded on the log.

#### Randomisation and Allocation Concealment

Allocation concealment will be maintained by use of a central, secure web randomisation process hosted at the University of Sydney’s Northern Clinical School Intensive Care Research Unit under the supervision of Associate Professor Gordon Doig. Blocks of variable size and a random seed will be used to ensure allocation concealment cannot be violated by deciphering the sequence near the end of each block. To further protect from deciphering, block size will not be revealed to site investigators. Randomisation is stratified by site. At randomisation each participant is assigned to nebulised heparin or standard care. There is a one to one allocation ratio.

#### Storage and Dispensing of Heparin for Nebulisation

Heparin sodium 25,000 Units in 5 mL manufactured by Pfizer Australia Pty Ltd (Sydney, Australia)^78^ or an equivalent unfractionated heparin by another approved manufacturer will be used. The medication will be stored in a secure area and dispensed by pharmacists and physicians under the supervision of investigators.

### Treatment of Participants

Participants assigned to ‘nebulised heparin’ will receive nebulised heparin in addition to the standard care required as determined by the treating team. Participants assigned to ‘standard care’ will receive the standard care required as determined by the treating team and will not be treated with nebulised heparin.

The nebulised heparin will be prescribed on the patient’s medication administration record by a treating physician. The prescription will specify the drug (unfractionated heparin sodium), the dose (25,000 Units in 5 mL), the frequency (6-hourly), the route (nebulised inhalation), the duration (10 days) and the indication (while receiving invasive ventilation in intensive care). DO NOT GIVE IV or SC.

Set-up of the nebuliser, ventilator circuit and expiratory filter is shown in **Figure 1**. Active ventilator circuit humidification will be used.

**Figure 1.**
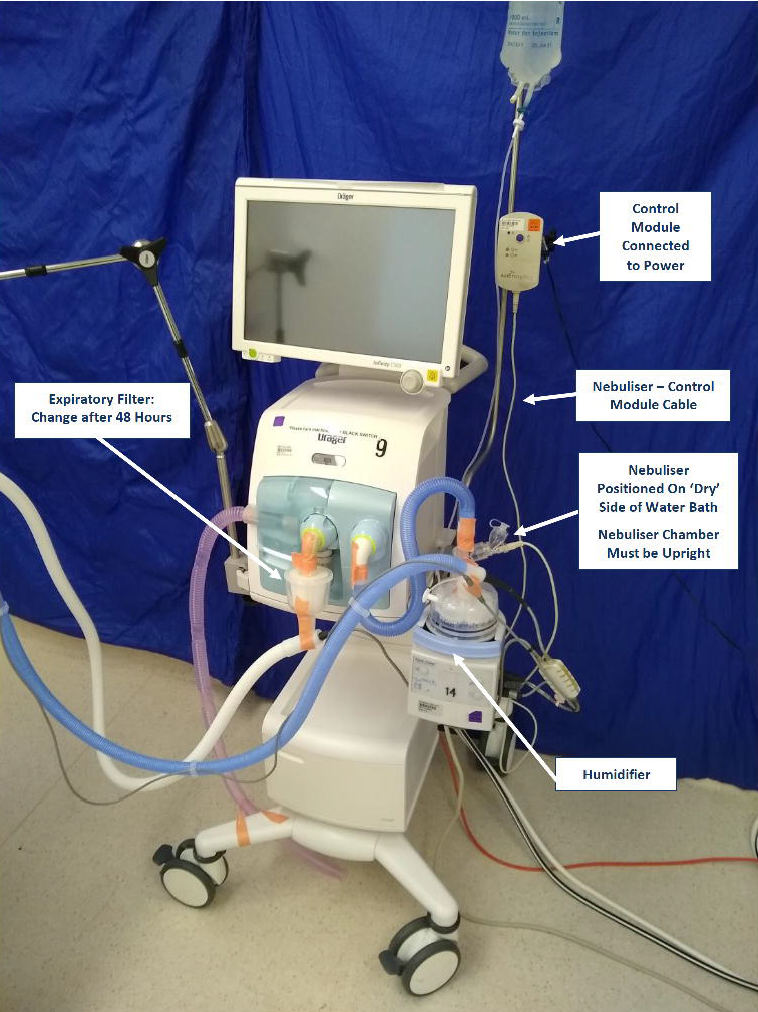
Ventilator circuit and nebuliser set-up

### Nebulisation

The Aerogen Solo^73,75,76^ vibrating mesh nebuliser is placed in the inspiratory limb of the ventilator circuit on the ‘dry’ side of the humidifier water bath. The nebuliser chamber must be upright.

An Aerogen nebuliser control module must be used. Some models of some ventilators have integrated (in-built) Aerogen nebuliser control modules. Examples of ventilators that might have integrated Aerogen nebuliser control modules are the Maquet SERVO-I, the GE Carescape and the Hamilton C6 and S1/G5. Using these integrated systems is acceptable. An Aerogen nebuliser control module that is external to the ventilator will be used if a ventilator-integrated (in-built) Aerogen nebuliser control module is not available.

### Expiratory filter

Some mechanical ventilators that incorporate an expiratory filter designed for use with nebulisation. If the ventilator does not have an internal expiratory filter, or if there is any doubt, a Servo Duo Guard (Maquet Critical Care AB)74 or an equivalent filter, will be placed at the end of the expiratory limb of the circuit and just before the expiratory valve of the ventilator. The filter is changed after 48 hours. The main purpose of the filter is to prevent exhaled nebulised heparin from depositing on the expiratory sensors and valves of the ventilator, but it is also a highly efficient viral filter that will mitigate against entry of exhaled virus into the room.

Each dose of nebulised heparin will be instilled into the nebuliser chamber using a sterile, single-use 5 mL syringe. The nebuliser chamber will be upright during nebulisation. The ventilator circuit will be inspected during nebulisation to verify that the medication is being delivered: it should be seen ‘misting’ in the circuit adjacent to the nebuliser chamber.

### Nebulising standard medications

The nebuliser used for heparin may also be used to administer preparations of salbutamol, ipratropium and budesonide that are approved for use by nebulisation.

Standard nebulised drugs should be scheduled so that the administration times of these drugs do not coincide with those of nebulised heparin.

Heparin should never be mixed with another drug or administered concurrently. Prior to instilling the heparin in the nebuliser chamber, the chamber must be inspected to ensure that the chamber is empty: drugs should never be mixed in the nebuliser chamber.

The nebuliser is very efficient and may deliver larger doses of standard nebulised drugs to the lungs compared to other nebulisers that are familiar to clinicians. The patient’s response to standard nebulised medications should be carefully monitored, especially if the recommended dose is exceeded.

### Withholding nebulised heparin

Treatment with any or all of the following therapies is not of itself reason to withhold nebulised heparin: deep vein thrombosis prophylaxis with unfractionated heparin or LMWH; ‘full’ therapeutic dose unfractionated heparin or LMWH; non-heparin anticoagulants; anti-thrombotic medications; protamine; prone positioning; and inhaled nitric oxide.

Nebulised heparin should be withheld if any of the following occurs:

- More than 10 days have elapsed since randomisation
- The patient is outside of ICU
- The patient is not receiving invasive ventilation
- The treating physician deems that there is a clinically unacceptable increase in APTT
- The treating physician deems that there is excessive bloodstaining of respiratory secretions
- There is pulmonary bleeding, major bleeding or suspected or confirmed HIT
- The patient is receiving ECMO or HFOV. Nebulised heparin should be recommenced if:
- Having been withheld because the patient was outside the ICU, the patient returns to ICU
- Having been withheld because the patient was not invasively ventilated, invasive ventilation is reinstituted
- Having been withheld because the APTT was unacceptably prolonged, the APTT becomes acceptable
- Having been withheld because there was excessive bloodstaining of respiratory secretions, the bloodstaining of the respiratory secretions has resolved
- Having been withheld for pulmonary bleeding or major bleeding, the bleeding is definitively controlled
- Having been withheld for suspected HIT, the patient is found not to have this condition
- Having been withheld for ECMO, the treatment with ECMO is stopped
- Having been withheld for HFOV, the treatment with HFOV is stopped.

## Data Collection

Paper records will be stored in locked rooms accessible only to authorised study personnel. Electronic information will be kept on password protected computers accessible only to authorised study personnel.

All study material, including case report forms and the study database, will be stored for a minimum period of 15 years after the conclusion of the study. Any paper study material that requires disposal will be shredded using a commercial grade shredder. Any electronic data requiring disposal will be thoroughly erased from its electronic media.

### Site master patient log

Each participating centre will maintain a log of enrolled patients that includes patient identifiers. Patient identifiers are not transferred to the study coordinating centre, but it must be possible to reidentify patients to allow future audit against source documents.

Case report form

Baseline data will be gathered from health records at the treating institution. It may also be necessary in a small number of cases to obtain information from other healthcare providers. The following baseline data is gathered:

- Eligibility criteria
- Intubation date and time; and intubation setting
- Birth date; sex; height; and weight
- History of tobacco smoking; hypertension; diabetes mellitus; asthma; and COPD
- Hospital admission date; Intensive care admission date and time; and APACHE III ICU diagnosis
- Treatment in the 24 hours prior to randomisation with unfractionated heparin, intravenously or subcutaneously; LMWH, intravenously or subcutaneously; lopinavir-ritonavir; remdesivir; hydroxychloroquine; interferon-β; interleukin antagonists; oseltamivir, laninamivir, zaninamivir or peramivir; macrolide; non-macrolide antibacterial; antifungal; and corticosteroid
- Treatment at the time of randomisation with inotrope or vasopressor infusion; renal replacement therapy; neuromuscular blocker; and prone positioning
- Serum creatinine, bilirubin and C-reactive protein; blood haemoglobin, white cell count and platelet count; and blood APTT, INR and D-dimer; and, for each, the result closest to and before randomisation
- For the chest radiograph (or chest CT) performed closest to and before randomisation: the number of lung quadrants with acute opacities that are not fully explained by effusions, lobar/lung collapse or nodules; whether the opacities are present bilaterally; whether, given all the medical information about the patient, the opacities are entirely attributable to cardiac failure or fluid overload; whether the patient was exposed to any risk factor for acute lung injury in the previous 7 days and; whether there is objective evidence to exclude the possibility of cardiac failure or fluid overload
- Arterial blood gas (pH, PaCO2, PaO2, bicarbonate and lactate) and corresponding ventilation parameters (respiratory rate, FIO2, peak airway pressure, PEEP or CPAP and tidal volume) closest to and before randomisation.

### Blood test results on days 1 to 10 while in intensive care

The following blood test results are gathered from health records at the treating institution for each day that the patient is in ICU on study days 1 to 10: lowest haemoglobin; highest white cell count; lowest platelet count; highest APTT; highest INR; highest D-dimer; and highest C-reactive protein.

Arterial blood gas results and ventilation observations on day 2 in intensive care Arterial blood gas (pH, PaCO2, PaO2, bicarbonate and lactate) and corresponding ventilation observations (respiratory rate, FIO2, peak airway pressure, PEEP or CPAP and tidal volume) closest to midday on day 2 are gathered from health records at the treating institution.

### Therapies on days 0 to 10 while in intensive care

The following data are gathered from health records at the treating institution for each day that the patient is in ICU up to and including day 10:

- Dose of nebulised unfractionated heparin
- Treatment with unfractionated heparin, intravenously or subcutaneously; LMWH, intravenously or subcutaneously; lopinavir-ritonavir; remdesivir; hydroxychloroquine; interferon-β; interleukin antagonists; oseltamivir, laninamivir, zaninamivir or peramivir; macrolide; non-macrolide antibacterial; antifungal; corticosteroid; inotrope or vasopressor infusion; renal replacement therapy; neuromuscular blocker; prone positioning; and extra-corporeal membrane oxygenation
- Volume of packed red cells and whole blood transfused.

### Day 28

The following data are gathered from health records at the treating institution and, where necessary, other healthcare providers:

- Date of first positive respiratory sample for SARS-CoV-2 and the sample type
- Date of first positive serology for SARS-CoV-2
- Acute hospital discharge status at day 28 and, if discharged, the date of discharge and discharge destination including deceased
- Readmission to ICU during the acute hospital admission and prior to day 28
- ICU status at day 28 and, if not in the ICU at the end of day 28, the final date and time of ICU discharge
- Invasive ventilation status at day 28 and, if not receiving invasive ventilation at the end of day 28, the final date and time that invasive ventilation was stopped
- Tracheotomy and, if performed, the procedure date.

### Day 60

The following data are gathered from health records at the recruiting institution and, where necessary, other healthcare providers and the patient or proxy:

- Vital status at day 60 and, if deceased, the date of death
- Place of residence at day 60
- If not discharged from the acute hospital discharge status by day 28, the acute hospital discharge status at day 60 is ascertained and, if now discharged, the date of discharge and the discharge destination including whether deceased.

### Adverse event or adverse reaction data

Adverse event or adverse reaction data are gathered from health records at the recruiting institution and, where necessary, other healthcare providers and the patient or proxy.

The following information is ascertained: the type of event or reaction, including with regard to pre-defined events (major bleeding, pulmonary bleeding and HIT); date and time of onset; date and time of most recent administration of nebulised heparin; the extent of any causal link to the study; the significance/severity of the event; action taken regarding the further use of nebulised heparin; and a freeform summary.

### The following adverse events and reactions must be reported

- Major bleeding which is bleeding that results in death and/or; bleeding that is symptomatic and occurs in a critical area or organ (intra-cranial, intra-spinal, intra-ocular, retroperitoneal, intra-articular, or intramuscular with compartment syndrome) and/or; bleeding that results in a fall in haemoglobin of 20g/L or more, or bleeding that results in transfusion of two or more units of whole blood or red cells
- Pulmonary bleeding, which is frank bleeding in the lungs, trachea or bronchi with repeated haemoptysis or requiring repeated suctioning and associated with acute deterioration in respiratory status
- HIT, which is an unexplained fall in platelet count and a positive heparin antibody test
- Adverse events and reactions that, in the site principal investigator’s judgement, are not part of the expected clinical course and could be related (at least possibly) to the study and are medically significant or had serious sequelae.

Blood transfusion is common in the study population, is recorded daily on the case report form up to day 10 and should not be reported as an adverse event unless the reason for transfusion is major bleeding or pulmonary bleeding.

Simple bloodstaining of the respiratory secretions is common in the study population and should not be reported as an adverse event.

Changes in APTT are common in the study population and should not be reported as an adverse event. The highest daily APTT is recorded on the case report form up to day 10. Period of vigilance for adverse reactions and events

Taking account of the pharmacodynamic profile of nebulised heparin 79, 80 and allowing a margin of safety, investigators should be alert to possible adverse events or reactions during the period from enrolment until 96 hours after the last dose of nebulised heparin.

#### Safety reporting process

Reportable adverse events and reactions will be communicated by site investigators to the chief investigator. In general, this will occur in writing within than 3 days of the site investigator becoming aware of the event. The management committee will assess all safety reports received from investigators. The chief investigator will provide the HREC and regulatory authorities with safety updates according to their reporting requirements.^81^

### Quality Assurance Monitoring

Conduct and progress of this trial will be monitored on an ongoing basis by the management committee. Case report forms received from the researchers will be assessed for protocol compliance. Remote data monitoring will be conducted by the management committee according to pre-defined criteria.

### Statistical Considerations

To demonstrate a clinically important 2.5-day improvement in the primary outcome, a sample size of 172 is required, assuming a decrease in day 28 mortality from 50% to 45% and a decrease in mean time to separation from invasive ventilation among day 28 survivors from 11 days to 8 days, with power 90% and a two-sided significance level of 0.05. To ensure safety, an interim analysis will be conducted after 60 patients have completed follow-up. Analyses will follow the intention-to-treat principle, considering all participants in the treatment group to which they were assigned, except for cases lost to follow-up or withdrawn. A detailed statistical plan will be published prior to database lock.

### Ethical Considerations

The study will be performed in accordance with the ethical principles of The Declaration of Helsinki,^82^ ICH Good Clinical Practice ^83^ and the National Statement on Ethical Conduct in Research Involving Humans.^84^ Approval of the protocol and related documents will be obtained from a HREC prior to the commencement of the study at each site. The investigators will ensure that all HREC conditions for the conduct of the study are met and that all requisite information is submitted to the responsible HREC.

## Data Availability

Data can be obtained by contacting the chief investigator

